# Low Seroprevalence of SARS-CoV-2 in Rhode Island Blood Donors Determined using Multiple Serological Assay Formats

**DOI:** 10.1101/2020.07.20.20157743

**Authors:** Daniel J. Nesbitt, Daniel Jin, Joseph W. Hogan, Philip A. Chan, Melissa J. Simon, Matthew Vargas, Ewa King, Richard C. Huard, Utpala Bandy, Christopher D. Hillyer, Larry L. Luchsinger

**Affiliations:** Lindsley F. Kimball Research Institute, New York Blood Center, New York, NY 10065, USA; Department of Biostatistics, Brown University, Providence, RI 02912, USA; Rhode Island Department of Health, Providence, RI 02906, USA; Rhode Island State Health Laboratory, Providence, RI, 02904, USA; Rhode Island Commerce Corporation, Providence, RI 02908, USA

## Abstract

Epidemic projections and public health policies addressing Coronavirus disease (COVID)-19 have been implemented without data reporting on the seroconversion of the population since scalable antibody testing has only recently become available. We measured the percentage of severe acute respiratory syndrome-Coronavirus-2 (SARS-CoV-2) seropositive individuals from 2,008 blood donors drawn in the state of Rhode Island (RI). We utilized multiple antibody testing platforms, including lateral flow immunoassays (LFAs), enzyme-linked immunosorbent assays (ELISAs) and high throughput serological assays (HTSAs). We report than an estimated seropositive rate of RI blood donors of approximately 0.6% existed in April-May of 2020. These data imply that seroconversion, and thus infection, is likely not widespread within this population. Daily new case rates peaked in RI in late April 2020. We conclude that IgG LFAs and HTSAs are suitable to conduct seroprevalence assays in random populations. More studies will be needed using validated serological tests to improve the precision and report the kinetic progression of seroprevalence estimates.

## Introduction

The Severe Acute Respiratory Syndrome Coronavirus (SARS-CoV)-2 pandemic is ongoing, with nearly 2.6 million cases and over 128,000 deaths reported from Coronavirus disease (COVID)-19 in the United States to date.^1,2^ Transmission models of SARS-CoV-2, based on numerous inferences of other immune responses to viral infections, suggest that infection may provide some immunity to reinfection.^1,3^ If true, the utility of serological tests to identify those who have acquired antibodies against SARS-CoV-2 (seroconversion) and the frequency of seroconversion in the population (seroprevalence) is a powerful tool with which to guide public health policies.^4,5^ It is critical to determine how many individuals have had COVID-19 and are thus likely to be immune, and differentiate them from those who have not been infected. These data are necessary to inform modeling projections and policy making that will allow an optimal approach to “reopening” a country, state, or region, and furthermore, these data must be accurate and reliable.

Serological assays rely on accurate recognition and ideally quantification of antibodies that recognize viral antigens specific to SARS-CoV-2. Optimal test characteristics include high levels of sensitivity and specificity. Coronaviruses have four major structural proteins; spike (S) protein (containing the S1 domain and RBD motif), nucleocapsid (N) protein, membrane (M) protein, and envelop (E) protein.^6^ Research conducted on 2005 SARS-CoV-1 and Middle East respiratory syndrome Coronavirus (MERS-CoV), which are highly related to SARS-CoV-2, found that recovered individuals produced the strongest immunogenic antibodies against antigens of the S- and N-proteins.^7^ Thus, the development of serological tests for SARS-CoV-2 antibodies has focused heavily on the detection of antibodies against these viral proteins. Antibody-based tests vary in both technology (platform) and target antigen (design). In May of 2020, the FDA announced a reversal in its emergency use authorization (EUA) and approval policies in order to help ensure that reliable tests are used to accurately measure seroconversion in a population. Some tests have received EUA but limited data is available. Considerable variability in test characteristics, particularly sensitivity, implies that there may not yet be an *ideal* test design and instrument platform. This also can lead to variability and potential bias in the estimation of the level of immunity in various locales or subpopulations.^8,9^

Multiple serological assays have been developed to detect SARS-CoV-2 antibodies from whole blood, plasma and serum. Essentially, three platforms of serological testing have been adopted: 1) in-house enzyme linked immunosorbent assays (ELISA), 2) high-throughput serological assays (HTSA) and 3) lateral flow assays (LFA). ELISAs offer wide flexibility for research laboratories to select virtually any antigenic protein of interest and assay patient sera to provide highly sensitive, quantitative results. HTSAs are more suitable to clinical laboratories processing large volumes of samples. Although HTSAs offer a narrower selection of antigen choices, these platforms offer high-throughput capacity, high sensitivity and can be integrated into clinical lab testing facilities. LFAs also offer limited antigen diversity, but function with small volumes of whole blood, plasma or sera (1 drop, ∼20uL) and require short test development times (≤30 minutes) allowing administration and test results at the point of care. As reagent supply, testing capacity and affordability vary across the country, the clinical community will undoubtedly resort to using multiple platforms to fill the demand.

Underreporting of COVID-19 cases may be occurring, which could inaccurately reflect the morbidity and mortality of SARS-CoV-2.^10^ The objective of this study was to assess the seroprevalence in a sample of blood donors in Rhode Island using commercially available serology tests .^11^ To this end, consecutive blood donors were enrolled though the Rhode Island Blood Center (RIBC) into a pilot study with the goal of estimating seroprevalence for the population represented by those who donate blood on a regular basis. This pilot is part of a larger statewide effort to estimate seroprevalence, including a statewide community survey and testing on specific populations of interest.

## Methods

### Whole Blood Donors and Sample Preparation

From April 27, 2020 – May 11, 2020, consecutive Rhode Island Blood Center (RIBC) donors (n=2,008) received a 2-question survey and completed a blood or plasma donation. Donor blood samples were then tested using two commercially available serology tests and an in-house ELISA, described below. Plasma or serum was isolated from whole blood samples collected in silica clot activator tubes. Samples were extracted, aliquoted to minimize future freeze-thaw cycles, and stored at -80°C.

### Lateral Flow ImmunoAssay (LFA)

LFAs were conducted using the Standard Q COVID-19 IgM/IgG Duo rapid immunochromatography test kit (SD Biosensor; South Korea).^12^ The kit contained two individual assay cartridges each with a detection band for IgG and IgM against SARS-CoV2 specific epitopes as well as an internal positive control. For each assay, 10 μL donor serum was applied to the sample pad, followed by two drops of proprietary running buffer according to the manufacturer’s instructions. After 15 min, a visual eye determination was made, and high-resolution images of the detection zone were taken and saved as .JPEG files. All tests were performed at room temperature.

### High-throughput Serology Assays

Serum samples were barcoded and dispatched to RIBC. Samples were analyzed using the VITROS Immunodiagnostic Products Anti-SARS-CoV-2 Total Ig Test with the VITROS 5600 (Ortho Clinical Diagnostics; USA). All assays were performed by trained RIBC employees according to the manufacturer’s standard procedures.

### In-house SARS-Cov2 Binding-Antibody ELISAs

Flat-well, nickel-coated 96 well ELISA plates (Thermo Scientific; USA) were coated with 2 ug/mL of recombinant S1 spike protein, nucleocapsid protein, or Receptor Binding Domain (RBD) spike protein specific to SARS-CoV-2 in resuspension buffer (1% Human Serum Albumin in 0.01% PBST) and incubated in a stationary humidified chamber overnight at 4°C. On the day of the assay, plates were blocked for 30 min with ELISA blocking buffer (3% W/V non-fat milk in PBST). Standard curves for both S1 and RBD assays were generated by using mouse anti-SARS-CoV spike protein monoclonal antibody (clone [3A2], ABIN2452119, Antibodies-Online) as the standard. Anti-SARS-CoV-2 Nucleocapsid mouse monoclonal antibody (clone [7E1B], bsm-41414M, Bioss Antibodies) was used as a standard for nucleocapsid binding assays. Monoclonal antibody standard curves and serial dilutions of donor sera were prepared in assay buffer (1% non-fat milk in PBST) and added to blocked plates in technical duplicate for 1 hr with orbital shaking at room temperature. Plates were then washed three times with PBST and incubated for 1 hr with ELISA assay buffer containing Goat anti-Human IgA, IgG, IgM (Heavy & Light Chain) Antibody-HRP (Cat. No. ABIN100792, Antibodies-Online) and Goat anti-Mouse IgG2b (Heavy Chain) Antibody-HRP (Cat. No. ABIN376251, Antibodies-Online) at 1:30000 and 1:3000 dilutions, respectively. Plates were then washed three times, developed with Pierce TMB substrate for 5 min, and quenched with 3 M HCl. Absorbance readings were collected at 450 nm. Standard curves were constructed in Prism 8.4 (Graphpad Software Inc.) using a Sigmoidal 4PL Non-Linear Regression (curve fit) model.

### Estimated Seroprevalence Statistical Calculations

For each assay, seroprevalence was estimated using a Bayesian statistical method that adjusts for sensitivity and specificity of the specific test. The operating characteristics for the Ortho assay were obtained from the technical report distributed by the manufacturer; for SD Biosensor we relied on local validation data. Details described in supplemental methods.

## Results

A total of 2,008 donor samples were collected for this study between April and May of 2020, just as the daily new case rates peaked in RI (https://ri-department-of-health-covid-19-data-rihealth.hub.arcgis.com/). We compared age, sex and race/ethnicity of the sample group to values reported for Rhode Island from the 2010 U.S. Census. The median age of donors was 56 years, significantly older than the Rhode Island median age of 39.4 years (**Fig. 1A, Table 1**). The sample had ∼47% female donors compared to 52% statewide (**Fig. 1B, Table 1**). The distribution of donor Race/ethnicity was 84.7% white, 2.7% Hispanic/Latino and 0.50% Black/African American, compared to the state distribution of 81% white, 12.4% Hispanic/Latino and 5.7% Black/African American. A full comparison appears in Table 1 and Figure 1. Notably, 9.3% of donors responded to ethnicity as ‘Declined’ or ‘Not Specified’. Finally, geographic location of donors associated with population density, such as Providence and Warwick, with lower representation in the western and coastal regions of Rhode Island (**Fig. 1C, 1D**). Thirteen donors were identified as convalescent plasma or whole blood donors that were aware of their seroconversion status prior to enrollment in the study and were removed from the analysis, which adjusted the total donors analyzed to 1,996.

**Table 1:** Distribution of Study Donor Age, Sex and Ethnicity compared to 2010 Rhode Island Population.

| Age Range | Study Donors | % Study Donors | RI Population | % RI Population |
| --- | --- | --- | --- | --- |
| 15-24 | 108 | 5.38% | 162,213 | 18.63% |
| 25-34 | 241 | 12.00% | 126,962 | 14.58% |
| 35-44 | 230 | 11.45% | 136,860 | 15.72% |
| 45-54 | 347 | 17.28% | 162,350 | 18.64% |
| 55-64 | 614 | 30.58% | 130,589 | 15.00% |
| 65-74 | 381 | 18.97% | 73,879 | 8.48% |
| 75+ | 87 | 4.33% | 78,002 | 8.96% |
| **Total** | 2,008 |  | 870,855 |  |

Source : http://www.dlt.ri.gov/lmi/census/demo/agesex.htm

| Gender | Study Donors | % Study Donors | RI Population | % RI Population |
| --- | --- | --- | --- | --- |
| Male | 1,064 | 53.01% | 508,400 | 48.30% |
| Female | 944 | 46.99% | 544,167 | 51.69% |
| **Total** | 2,008 |  | 1,052,567 |  |

Source : http://www.dlt.ri.gov/lmi/census/demo/agesex.htm

| Ethnicity | Study Donors | % Study Donors | RI Population | % RI Population |
| --- | --- | --- | --- | --- |
| White | 1,700 | 84.66% | 856,869 | 81.41% |
| Black or African American | 10 | 0.50% | 601,89 | 5.72% |
| American Indian & AK Native | 5 | 0.25% | 6,058 | 0.58% |
| Asian | 5 | 0.25% | 30,457 | 2.89% |
| Hawaiian/Pacific Islander | 0 | 0.00% | 554 | 0.05% |
| Hispanic/Latino or Other Race, Alone | 59 | 2.94% | 63,653 | 6.05% |
| Two or More Races | 20 | 1.00% | 34,787 | 3.30% |
| Decline | 23 | 1.154% | 0 | 0.00% |
| Not Specified | 186 | 9.26% | 0 | 0.00% |
| **Total** | 2,008 |  | 870,855 |  |

Source : http://www.dlt.ri.gov/lmi/census/demo/ethnic.htm

**Figure 1:**
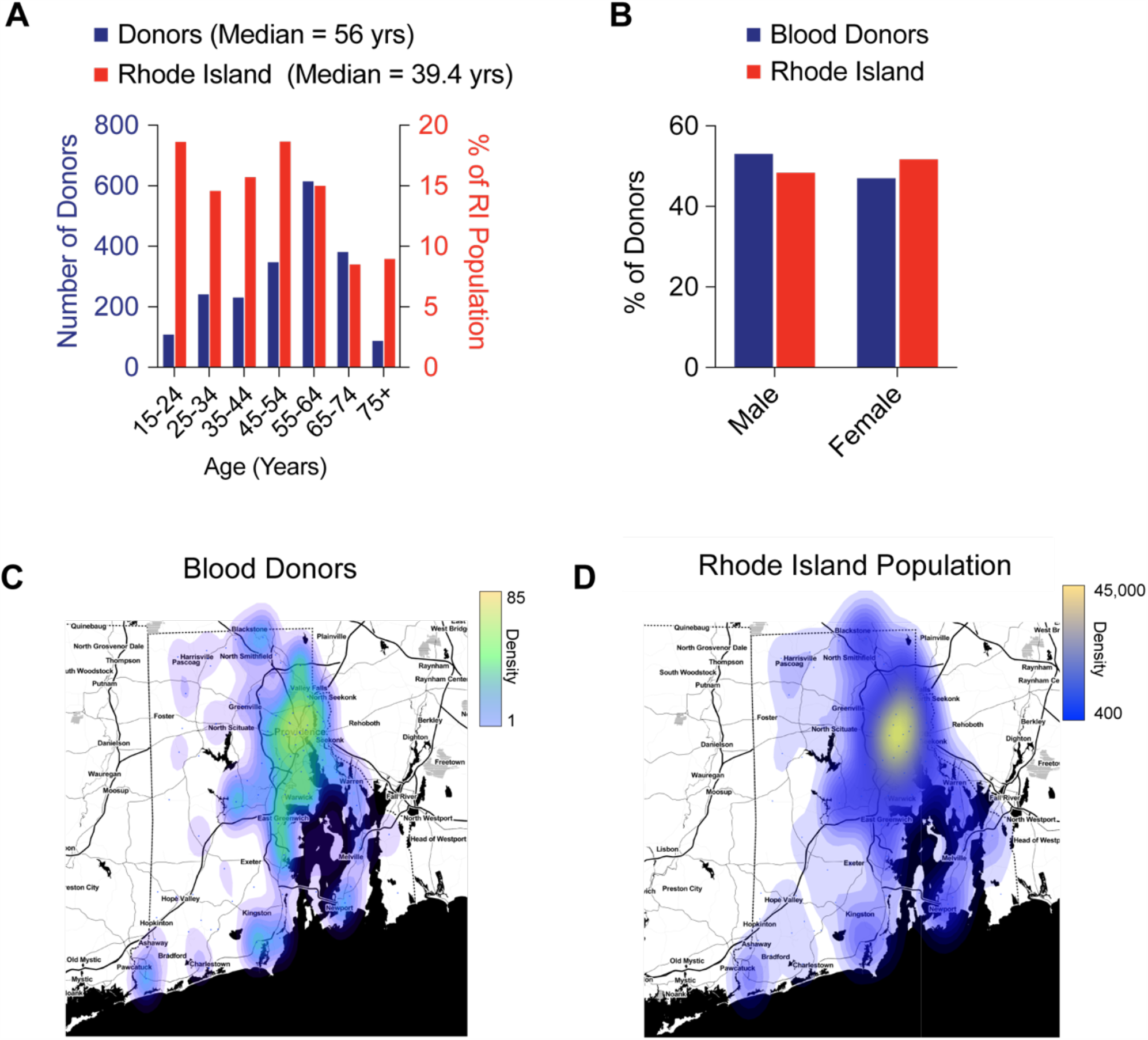
Demographics of Rhode Island Seroprevalence Donors. **A;** Distribution of seroprevalence donor age (blue bars) compared to RI population (red bars). N=2008 **B;** Distribution of seroprevalence donor sex (blue bars) compared to RI population (red bars). N=2008. **C;** Choropleth of zip codes for all seroprevalence blood donors. **D;** Choropleth of zip codes for RI population (right).

To quantify seroprevalence in this sample, donor samples were tested with an HTSA platform (Ortho Clinical Diagnostics VITROS Total Ig Test) and an LFA platform (SD Biosensor IgM/IgG test). The IgM-only LFA assay yielded 68 positive tests for a 2.7% (95% CI 1.7 to 3.8%) seroconversion (**Fig. 2A, Table 2**). In contrast, the IgG-only LFA assay yielded 13 positive tests for 0.6% seroconversion (95% CI 0.3 to 1.1%) and was in agreement with the Ortho HTSA assay, which had 14 positives for a 0.6% seropositivity (95% CI 0.2 to 1.1) (**Fig. 2A, Table 2**).

**Table 2.**
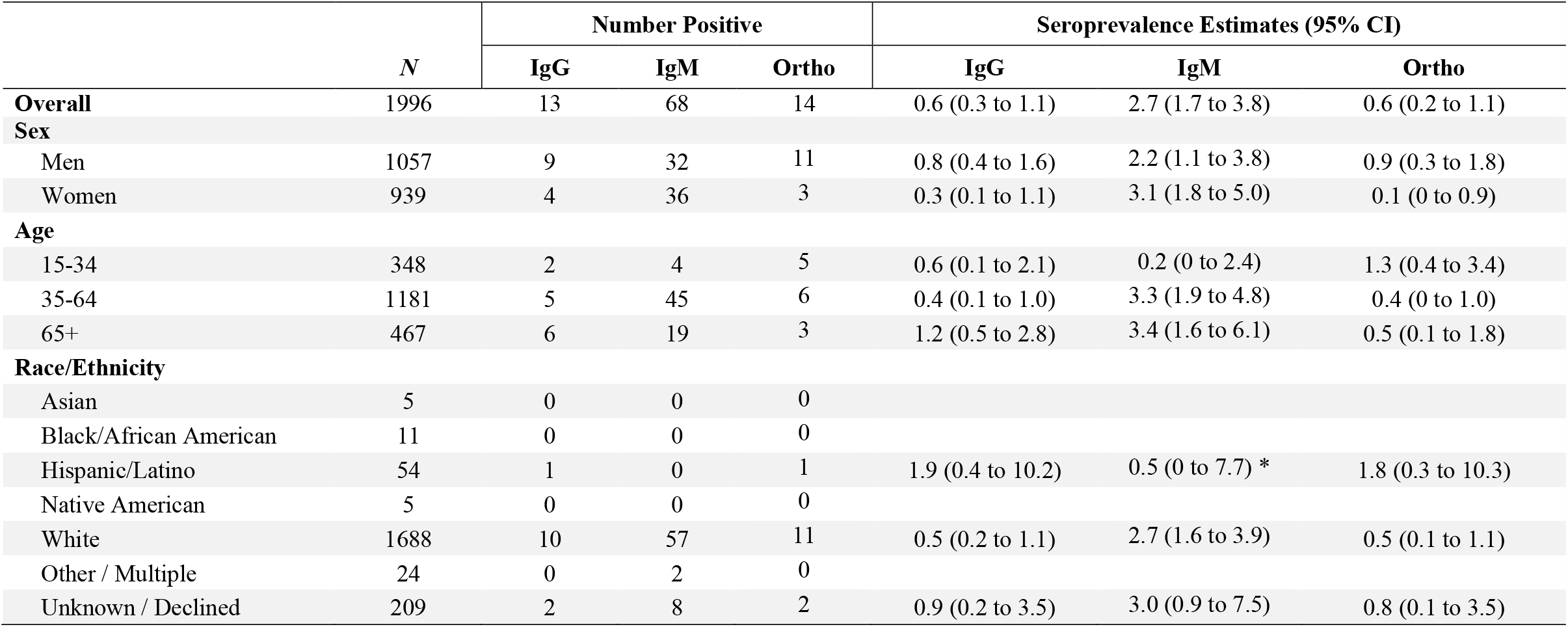
Antibody test results and seroprevalence estimates overall and by sex, age and race/ethnicity. Seroprevalence estimates reported in terms of posterior mode and 95% credible interval, calculated using Bayesian method that adjusts for test sensitivity and specificity. Estimates not reported for categories with 25 test results or fewer. Excludes 11 positive CP/WB Donors and 2 CP/WB donors that tested negative for all three tests *Posterior mode calculated using a prior distribution having mode equal to the overall seroprevalence for IgM

**Figure 2.**
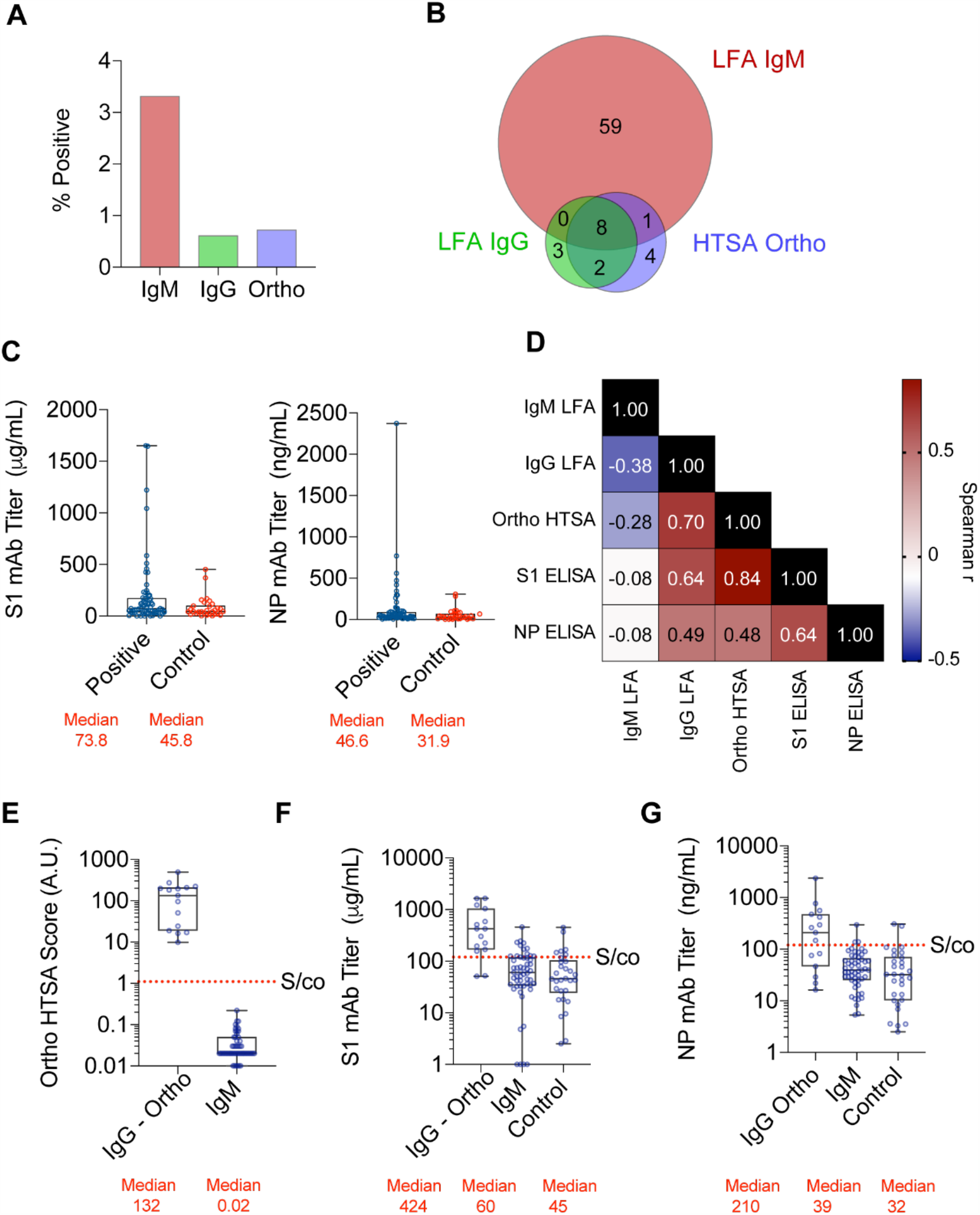
**A;** Percent of donors testing positive using IgM LFA (red), IgG LFA (green) or Ortho HTSA (blue). **B;** Venn diagram of seropositive samples using IgM LFA, IgG LFA and Ortho HTSA. **C;** Monoclonal antibody quantification of all seropositive samples using S1 spike protein (left) and Nucleocapsid (N) protein (right) ELISA assays. Median values are indicated in red. **D;** Spearman correlation coefficients, r, of serological assays. N=77 samples. **E-G;** Serological results of IgG-Ortho and IgM-only seropositive groups using Ortho HTSA (left), S1 ELISA (center) and NP ELISA (right) assays. Signal to cutoff (S/co) for each assay is indicated.

In total, 3.9% of all samples (77 seropositive donors) were reactive for at least one test. To report overlap between test results, we constructed a Venn diagram (**Fig 2B, Table 2**). Notably, ∼76% of seropositive samples (59 of 77) were reactive only with the IgM-only LFA test. The remaining 0.9% of all samples (18 seropositive donors) showed a ∼62% overlap between Ortho and IgG LFA assays (10 of 18 seropositive donors). Samples that showed at least 2 or more positive reactions was 0.55% (11 seropositive donors).

Donors completed a two-part questionnaire as to whether they had COVID-19 and if so, the results of the diagnostic PCR test. Overall, 76 donors responded that they had received a diagnostic PCR test for COVID-19; of these, 13 donors tested positive while 63 tested negative (**Table 3**). Of those reporting positive PCR, 4/13 (44%) had positive IgM LFA, 9/13 (69%) had positive IgG, and 11/13 (85%) had positive Ortho test. Of those reporting negative PCR, 59/63 (94%) tested IgM negative, 61/63 (97%) tested IgG negative, and the same number (97%) tested Ortho negative. These limited data are in line with manufacturer-reported estimates of sensitivity and specificity. Importantly, the reliance on self-reported data must be interpreted with caution, and there was no ability to account for the time since infection, which could impact the sensitivity calculations.

**Table 3:** Serology Test Results stratified by reported PCR test result among SARS-CoV-2 Diagnostic PCR Test Respondents.

|  |  | PCR Result |  | Total |
| --- | --- | --- | --- | --- |
|  |  | Positive | Negative |  |
| IgM | Positive | 4 | 4 | 8 |
|  | Negative | 9 | 59 | 68 |
|  | Total | 13 | 63 | 76 |
|  |  | PCR Result |  | Total |
|  |  | Positive | Negative |  |
| IgG | Positive | 9 | 2 | 11 |
|  | Negative | 4 | 61 | 65 |
|  | Total | 13 | 63 | 76 |
|  |  | PCR Result |  | Total |
|  |  | Positive | Negative |  |
| Ortho | Positive | 11 | 2 | 13 |
|  | Negative | 2 | 61 | 63 |
|  | Total | 13 | 63 | 76 |

The gold-standard in antibody quantification is the ELISA assay for its flexibility in antigen diversity and quantification methodology using monoclonal antibodies to generate standard curves. We designed in-house ELISA assays against S1 and NP specific to SARS-CoV-2 antibodies, since these antigens have been described to elicit the most immunogenic response to infection based on SARS-CoV and MERS research. We analyzed all 77 samples that were positive for any serological assay and 30 random samples that were negative for all serological assays as controls for S1 and NP antibodies. Surprisingly, S1 antibody quantification showed a median value of 73.8μg/mL for seropositive samples compared to 45.8μg/mL for seronegative controls (Fig. 2C) indicating moderate antibodies against S1 epitopes. Similarly, NP antibody quantification showed a median value of 46.6ng/mL for seropositive samples compared to 31.9ng/mL for seronegative controls, also indicating moderate antibodies against NP epitopes. However, there was ≥100-fold range of antibody values for seropositive samples in each ELISA test, suggesting that some of the seropositive samples, but not all, were significantly reactive in S1 and NP ELISA, which is highly predictive of neutralizing activity. Correlation analysis of all five tests showed a high degree of positive association between ELISA, HTSAs and IgG LFA tests while IgM LFA test was negatively correlated (Fig. 2E). Thus, we hypothesized that samples reactive for either IgG LFA and/or Ortho HTSA may have higher ELISAs values than samples that were reactive only for IgM LFA test.

To investigate this, we subdivided seropositive samples into “IgG/Ortho” or “IgM-only” groups. As expected, the median Ortho HTSA value was 10^4^ higher for the IgG/Ortho group than the IgM-only group (134A.U. vs 0.02A.U., respectively) (**Fig. 2E**). Similarly, both S1 and NP ELISAs showed significantly higher median antibody concentrations for the IgG/Ortho group than for the IgM-only group (S1; 424.2μg/mL versus 60.5μg/mL and NP; 210.1ng/mL versus 39.4ng/mL) (**Fig. 2F, 2G**). Importantly, these results conclude that IgG LFA and Ortho HTSA assays, but not the IgM LFA assay, correlate with immunogenic antibodies specific to SARS-CoV-2 as detected by ELISA.

## Discussion

This is among the first studies to evaluate statewide seroprevalence using blood donations. COVID-19 antibody testing has entered public discourse as an important metric in determining the population seroprevalence of SARS-CoV-2. Ultimately, the application of antibody testing could be clinically informative as to the degree of immunity afforded incurred by recovered patients or to that of future vaccinated individuals. However, we recognize the limitations of the current study include generalizability and limited demographic and other data of the blood donors that may be important. In fact, seroprevalence has been suggested to be higher in specific racial/ethnic communities based on recent studies.^13^ Thus, more inclusive and complete seroprevalence studies will need to be performed in the future.

The application of antibody testing could be clinically informative as to the degree of antiviral activity incurred by recovered patients or to that of future vaccinated individuals. Seroprevalence studies have the ability to provide two important metrics: 1) the seroprevalence within a given population and 2) semi-quantification of specific antibodies to SARS-CoV-2 that may correlate with immunity. However, the latter estimation requires that an accurate methodology be adopted at the onset of the study. We recently completed a comprehensive analysis of SARS-CoV-2 serological test characteristics and comparison to antiviral neutralization activity using pseudoviral models.^14^ In that investigation, HTSAs were shown to have superior performance characteristics and correlation with neutralizing activity compared to LFAs. It should be noted that the LFAs used in the prior study were different from the LFAs used in this study.

Among Rhode Island blood donors, we found the SD Biosensor IgG LFA and the Ortho HTSA assays both reported a ∼0.6% estimated seroprevalence rate. This is in agreement with a recent study showing relatively low seroprevalence in many metropolitan areas.^15^ It is tempting to speculate that low rates of seroprevalence is a logical result to the social distancing and mitigation policies that have been adopted by virtually the entire world. However, the SD Biosensor IgM LFA assay had very different performance characteristics, did not correlate with ELISA assays and reported a higher seroprevalence rate. The latter approximation would be similar to the Santa Clara seroprevalence rate reported in April of 2020, which found a seroprevalence rate of 2.5-4.2% using LFA assays.^16^ However, since the IgM LFA assay correlated poorly with the Ortho HTSA assay, which we have previously shown to associate with neutralization activity and antiviral antibody effectiveness to prevent reinfection of cells with pseudovirus, ^14^ we conclude the SD Biosensor IgM LFA assay is not informative as to a specific adaptive immune response to SARS-CoV-2. It should be noted that a concurrent SARS-CoV-2 serology study comparing the SD Biosensor LFAs to another LFA and a chemiluminescent assay concluded that the SD Biosensor IgM LFA had limited clinical utility, while the SD Biosensor IgG LFA performed very well across several distinct population sets and compared to the other assays (Dr. Shaolei Lu et al.; manuscript submitted). Our results caution that seroprevalence rates could be miscalculated by as much as 5-fold depending on the type of serology test employed. Only assays that show significant correlation to neutralization activity, a metric of specific adaptive immunity, should be employed to report rates of seroprevalence.

LFAs offer the convenience of rapid test results at the point of care and utilization of either whole blood, plasma or serum which makes deployment simple. In this study, we found that the SD Biosensor IgG LFA test provided reliable sensitivity to report seroprevalence. We found in this study that the SD Biosensor IgG LFA test also provided reliable sensitivity to report seroprevalence. However, LFAs do not yield semi-quantitative results which could be used to further understand the immunological range of responses within a study population. Therefore, HTSA platforms are better suited to quantify a wide range of antibody levels in a population while LFAs are suitable for low-cost, rural or studies designed for a limited interpretation of seroprevalence.

In conclusion, we find the estimated seroprevalence of Rhode Island blood donors to be relatively low, approximately 0.6%. Thus, we predict undiagnosed and asymptomatic infections are also likely to be low. Considering the possibility that this may be an underestimate of the statewide population, these conclusions draw important findings as it suggests that in the absence of a vaccine, “background” or “herd” immunity to also be low, now four months into the US pandemic, and thus the susceptible population remains at 95% or greater.

## Data Availability

All data is available in the manuscript.

## References

1 Andersen, K. G., Rambaut, A., Lipkin, W. I., Holmes, E. C. & Garry, R. F. The proximal origin of SARS-CoV-2. Nat Med 26, 450–452, doi:10.1038/s41591-020-0820-9 (2020).

2 Wu, F. et al. A new coronavirus associated with human respiratory disease in China. Nature 579, 265–269, doi:10.1038/s41586-020-2008-3 (2020).

3 Kucharski, A. J. et al. Early dynamics of transmission and control of COVID-19: a mathematical modelling study. Lancet Infect Dis 20, 553–558, doi:10.1016/S1473-3099(20)30144-4 (2020).

4 Chan, C. M. et al. Examination of seroprevalence of coronavirus HKU1 infection with S protein-based ELISA and neutralization assay against viral spike pseudotyped virus. J Clin Virol 45, 54–60, doi:10.1016/j.jcv.2009.02.011 (2009).

5 Lee, C. Y., Lin, R. T. P., Renia, L. & Ng, L. F. P. Serological Approaches for COVID-19: Epidemiologic Perspective on Surveillance and Control. Front Immunol 11, 879, doi:10.3389/fimmu.2020.00879 (2020).

6 Zhong, X. et al. B-cell responses in patients who have recovered from severe acute respiratory syndrome target a dominant site in the S2 domain of the surface spike glycoprotein. J Virol 79, 3401–3408, doi:10.1128/JVI.79.6.3401-3408.2005 (2005).

7 Tai, W. et al. Characterization of the receptor-binding domain (RBD) of 2019 novel coronavirus: implication for development of RBD protein as a viral attachment inhibitor and vaccine. Cell Mol Immunol, doi:10.1038/s41423-020-0400-4 (2020).

8 Chaturvedi, R., Naidu, R., Sheth, S. & Chakravarthy, K. Efficacy of Serology Testing in Predicting Reinfection in Patients with SARS-CoV-2. Disaster Med Public Health Prep, 1–7, doi:10.1017/dmp.2020.216 (2020).

9 Deeks, J. J. et al. Antibody tests for identification of current and past infection with SARS-CoV-2. Cochrane Database Syst Rev 6, CD013652, doi:10.1002/14651858.CD013652 (2020).

10 Lachmann, A. Correcting under-reported COVID-19 case numbers. medRxiv, doi:10.1101/2020.03.14.20036178 (2020).

11 Xu, X. et al. Seroprevalence of immunoglobulin M and G antibodies against SARS-CoV-2 in China. Nat Med, doi:10.1038/s41591-020-0949-6 (2020).

12 Kimberly J Paiva, R. D. G., Philip A Chan, John R. Lonks, Ewa King, Richard C Huard, Diane L Pytel-Parenteau, Ga Hie Nam, Evgeny Yakirevich, Shaolei Lu. Validation and Performance Comparison of Three SARS-CoV-2 Antibody Assays. bioRxiv, doi:10.1101/2020.05.29.124776 (2020).

13 Martinez, D. A. et al. SARS-CoV-2 Positivity Rate for Latinos in the Baltimore-Washington, DC Region. JAMA, doi:10.1001/jama.2020.11374 (2020).

14 Luchsinger, L. L. et al. Serological Analysis of New York City COVID19 Convalescent Plasma Donors. medRxiv, doi:10.1101/2020.06.08.20124792 (2020).

15 Havers, F. P. et al. Seroprevalence of Antibodies to SARS-CoV-2 in Six Sites in the United States, March 23-May 3, 2020. medRxiv, doi:10.1101/2020.06.25.20140384 (2020).

16 Bendavid, E. et al. COVID-19 Antibody Seroprevalence in Santa Clara County, California. doi:10.1101/2020.04.14.20062463 (2020).

